# Early IFNβ secretion determines variable downstream IL-12p70 responses upon TLR4 activation in health and disease

**DOI:** 10.1101/2020.12.19.20248132

**Authors:** Celine Posseme, Alba Llibre, Bruno Charbit, Vincent Bondet, Vincent Rouilly, Violaine Saint-André, Jeremy Boussier, Jacob Bergstedt, Nikaïa Smith, Maxime Rotival, Michael S Kobor, Tom Scriba, Estelle Mottez, Stanislas Pol, Etienne Patin, Matthew L. Albert, Lluis Quintana-Murci, Darragh Duffy, Milieu Intérieur Consortium

**Affiliations:** Translational Immunology Lab, Institut Pasteur, Paris, France; Frontière de l’Innovation en Recherche et Education (FIRE) doctoral school, Paris, France; Cytometry and Biomarkers UTechS, CRT, Institut Pasteur, Paris, France; DATACTIX, Paris, France; Bioinformatics and Biostatistics HUB, Institut Pasteur, USR 3756 CNRS, Paris, France; Human Evolutionary Genetics Unit, Institut Pasteur, UMR2000, CNRS, Paris 75015, France; Department of Medical Genetics, Center for Molecular Medicine and Therapeutics, University of British Columbia/British Columbia Children’s Hospital Research Institute, Vancouver, BC, Canada; South African Tuberculosis Vaccine Initiative (SATVI), Division of Immunology, Department of Pathology and Institute of Infectious Disease and Molecular Medicine, University of Cape Town, South Africa; Hepatology Unit, Hôpital Cochin, AP-HP, 27, rue du Fg Saint-Jacques, 75014 Paris, France; Insitro, San Francisco, California, USA; Human Genomics and Evolution, Collège de France, Paris 75005, France

**Keywords:** Cytokines, IL-12p70, IFNβ, systems immunology, immune variability, HCV

## Abstract

The IL-12 family of cytokines comprises the only heterodimeric cytokines mediating diverse functional effects. We previously observed a bi-modal IL-12p70 response to LPS in healthy donors of the *Milieu Interieur* cohort. Herein, we demonstrate that IFNβ expression serves as an upstream determinant of variable IL-12p70 production. Integrative modelling of proteomic, genetic, epigenomic and cellular data confirmed IFNβ as key for regulation of LPS induced *IL12A* and IL-12p70 variability. The clinical relevance was supported by reduced and variable IL-12p70 responses in individuals infected with the hepatitis C virus (HCV), and findings that IFN-based therapy for HCV is more likely to fail in those patients with dysregulated pre-treatment IL-12p70 responses. In sum, our systems immunology approach has defined a better understanding of IL-12p70 and IFNβ in healthy and infected persons, providing insights into how common genetic and epigenetic variation may impact immune responses to bacterial infection in health and disease.

## Introduction

Upon infection, a balanced immune response is crucial to respond to the pathogen while maintaining homeostasis. Insufficient immune responses can increase susceptibility to infections, while an overactive immune system may lead to autoimmune disorders such as type 1 diabetes or lupus (Atkinson et al., 2014) (Tsokos et al., 2016). Although cytokines are essential regulators of both homeostasis and inflammatory responses, few studies have been conducted in humans to understand the causes of variance in healthy individuals before applying this understanding to disease cohorts (Ter Horst et al., 2016) (Li et al., 2016b) (Brodin et al., 2015) (Enroth et al., 2014). We hypothesized that understanding the underlying reasons of how variance exists will provide new fundamental understanding in disease states, with potential clinical relevance.

A crucial cytokine for response to infection is IL-12p70, a heterodimeric pro-inflammatory cytokine composed of the p35 and p40 subunits, which induces Th1 responses (Kobayashi et al., 1989). IL-12p70 induces the proliferation and cytotoxicity of NK cells as well as the differentiation and proliferation of Th1 cells, primarily through the induction of IFNγ and subsequent Jak/STAT signaling (Trinchieri, 2003). Although p35 requires binding to p40 to be secreted (Wolf et al., 1991), p40 can function autonomously as a monomer (IL-12p40) or homodimer (IL-12p80), or it can bind to the p19 subunit to form the pro-inflammatory cytokine IL-23 (Ling et al., 1995) (Oppmann et al., 2000). IL-12p40 can antagonize IL-12p70-driven responses by competing for the receptor, whereas IL-23 facilitates Th17 responses (Vignali and Kuchroo, 2012). Given this complexity, a better understanding of the factors that determine the variable functions of IL-12 cytokines may have broad implications for understanding diverse immune functions in homeostasis and disease.

We previously reported high levels of inter-individual variability in IL-12p70 responses to LPS stimulation in a small cohort of healthy donors (Duffy et al., 2014). Here, we dissected the factors behind this variability in 1,000 healthy individuals of the *Milieu Interieur* (MI) cohort. Using standardized immunomonitoring tools, we identified that a quarter of healthy individuals consistently secreted low levels of IL-12p70 cytokine after lipopolysaccharide (LPS) stimulation. We applied a systems biological approach integrating proteomic, transcriptomic, cellular, epigenetic, and genomic data sets to assess environmental and intrinsic factors driving IL-12p70 variability. From this, we identified upstream variability in IFNβ as the major determinant of variable IL-12p70 responses. Supporting the potential clinical relevance of these findings we show that the IFNβ-IL-12p70 pathway was perturbed in chronically infected HCV patients that failed IFN based treatment. Our study adds new understanding to IL-12p70 responses in a healthy population and provides a model for dissection of variable cytokine responses for new insights into disease pathogenesis.

## Results

### TLR4 activation induces variable and stable IL-12p70 responses in a healthy population

We initially measured the secretion of 32 proteins following stimulation of whole-blood with 27 unique stimuli and observed high levels of inter-individual variability in IL-12p70 cytokine responses to LPS in 25 healthy donors of the MI cohort (Duffy et al., 2014). To further characterize potential reasons behind this cytokine variability, we examined IL-12p70 expression by Luminex assay in 1,000 healthy donors of the MI cohort, for whom whole blood was stimulated for 22 hours with LPS (TLR4 agonist), Poly(I:C) (TLR3 agonist), or left unstimulated (null control). Although an IL-12p70 response was detected for all donors upon TLR3 activation, 28% of healthy donors did not secret detectable levels of IL-12p70 upon TLR4 stimulation, suggesting that the variation observed for IL-12p70 is specific to the TLR4 induced response (**Fig 1A**). As IL-12p70 is a heterodimer molecule composed of IL-12p35 and IL-12p40, the latter also forming IL-23 (with IL-12p19), we examined whether these responses showed similar variability after LPS stimulation. Measurement of IL-12p40 (**Fig 1B**) and IL-23 (**Fig 1C**) in LPS-stimulated supernatants revealed a normal distribution of responses suggesting that the observed variability was specific for the IL-12p70 heterodimer cytokine.

**Figure 1.**
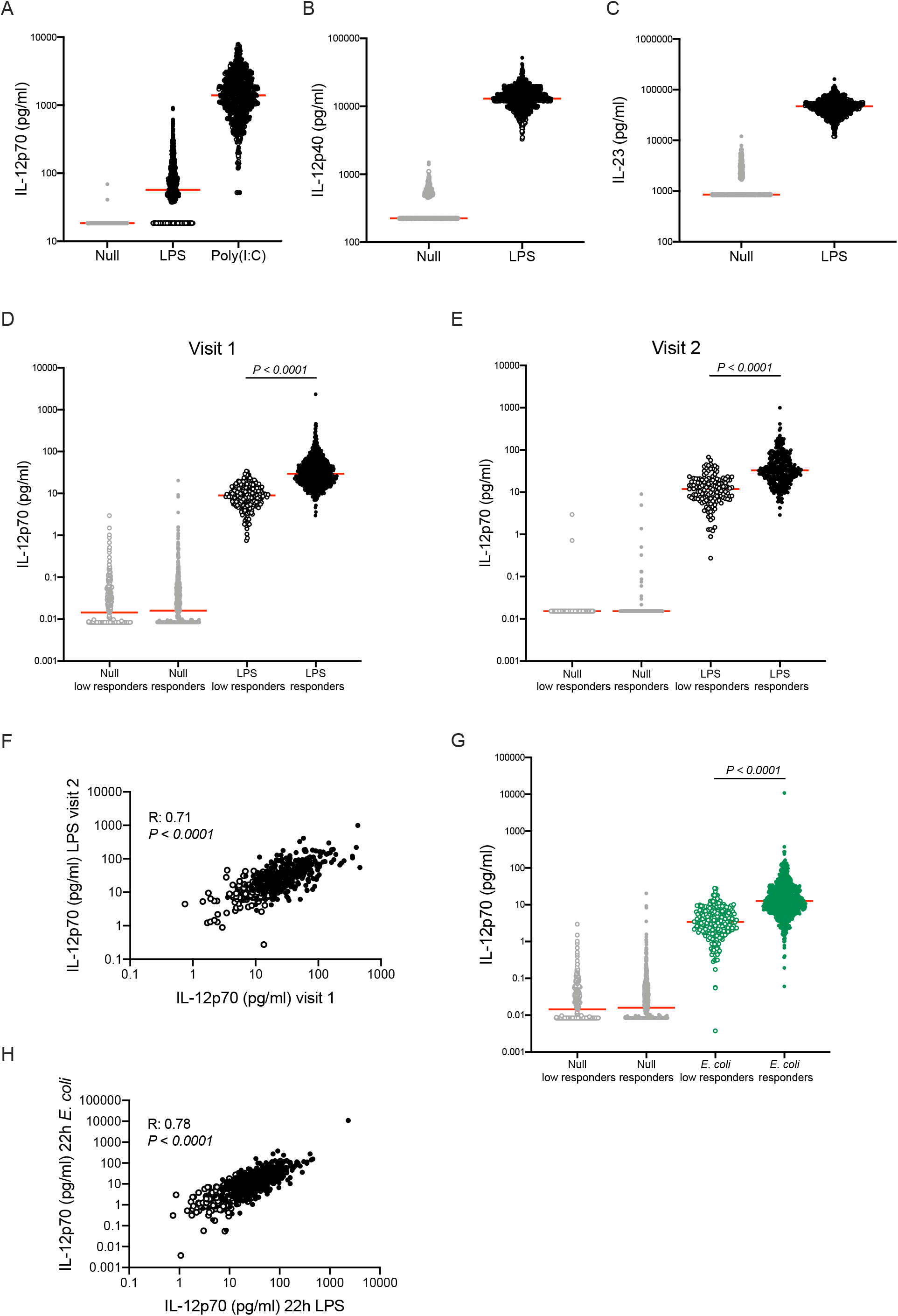
IL-12p70 variability in healthy donors. **(A)** IL-12p70 measured by Luminex in healthy donors (n=1,000) after stimulation of whole blood with null, LPS, or Poly(I:C). (**B**) IL-12p40 and (**C**) IL-23 proteins measured by Luminex in healthy donors (n=1,000) after null and LPS stimulation. IL-12p70 secretion measured by Simoa after null and LPS stimulation at two separate time points (Visit 1 n=1,000 (**D**); Visit 2 n=500 (**E**)). (**F**) Correlation of IL-12p70 secretion after LPS 22h stimulation at 2 separate time points. Pearson correlation was performed on log10 values. (**G**) IL-12p70 secretion measured by Simoa in the two groups of *Milieu Interieur* IL-12p70 responders after null and *E. coli* stimulation. Red lines indicate the median value for each group. P values were determined by an unpaired Student’s t test using log10 values. (**H**) Correlation of IL-12p70 secretion measured by Simoa after *E. coli* and LPS 22h stimulation. Pearson correlation was performed using log10 values. Empty symbols represent IL-12p70 low responders as defined by LPS 22 hours whole blood stimulation (1A), full symbols represent IL-12p70 responders.

To test the hypothesis that the 28% of MI donors secreted IL-12p70 concentrations below the detection limit of the Luminex assay we developed an ultrasensitive Simoa digital ELISA. This novel assay has a limit of detection (LoD) of 0.01 pg/ml, which is 1000 times more sensitive than Luminex (LoD: 18.5 pg/ml). Using this assay, we detected IL-12p70 secretion from all donors, and therefore re-defined the 28% of donors that had undetectable levels of IL-12p70 with Luminex, as low responders. As a group these low responders had significantly lower levels of IL-12p70 as compared to the responders (*P* < 0.0001) (**Fig 1D**). To test if this phenotype was reproducible, we measured IL-12p70 in 500 MI donors who were sampled at a second time point between 2 to 6 weeks later (Visit 2). The IL-12p70 response at Visit 2 was also significantly different (*P* < 0.0001) between the two groups as observed in Visit 1 (**Fig 1E**), and the IL-12p70 responses over the 2 timepoints showed a strong and significant correlation (R = 0.71, *P* < 0.0001) (**Fig 1F**).

We also confirmed that this difference was present after *E. coli* stimulation (**Fig 1G**), and that the IL-12p70 variance for LPS and *E. coli* (R = 0.78, *P* < 0.0001) was consistent (**Fig 1H**). Overall, these results indicate that inherent variability in the IL-12p70 response after TLR4 activation is a consistent and prevalent phenotype.

### IL-12p70 low responders have fewer activated monocytes and dendritic cells

We next asked whether this consistently lower IL-12p70 response was due to differences in circulating immune cells. To first define the immune cells that produce IL-12p70, we stimulated whole blood of 10 healthy donors with LPS for 22 hours in the presence of a protein transport inhibitor (added 8 hours after stimulation initiation). A combination of 10 surface antibodies was used to characterize the major circulating immune cell populations (**Table S1**). As IL-23 shares the p40 subunit with IL-12p70, both IL-12p70 and IL-23 were assessed intracellularly in 8 relevant cell types. The null condition was used to determine the positivity threshold for IL-12p70 and IL-23 secretion (**Fig 2A**). We observed that CD14^+^ monocytes and dendritic cells (DCs) were the main producers of both IL-12p70 and IL-23 cytokines upon LPS stimulation (Wilcoxon test, Q value < 0.05) (**Fig 2B** and **Fig 2C**). Interestingly, more than 30% of CD14^+^ monocytes secreted both IL-12p70 and IL-23, whereas around 7% of CD14^+^ monocytes secreted IL-12p70 only. A similar observation was made with conventional dendritic cells 2 (cDC2) (**Fig 2D**). Thus, the majority of cells producing IL-12p70 also secrete IL-23.

**Figure 2.**
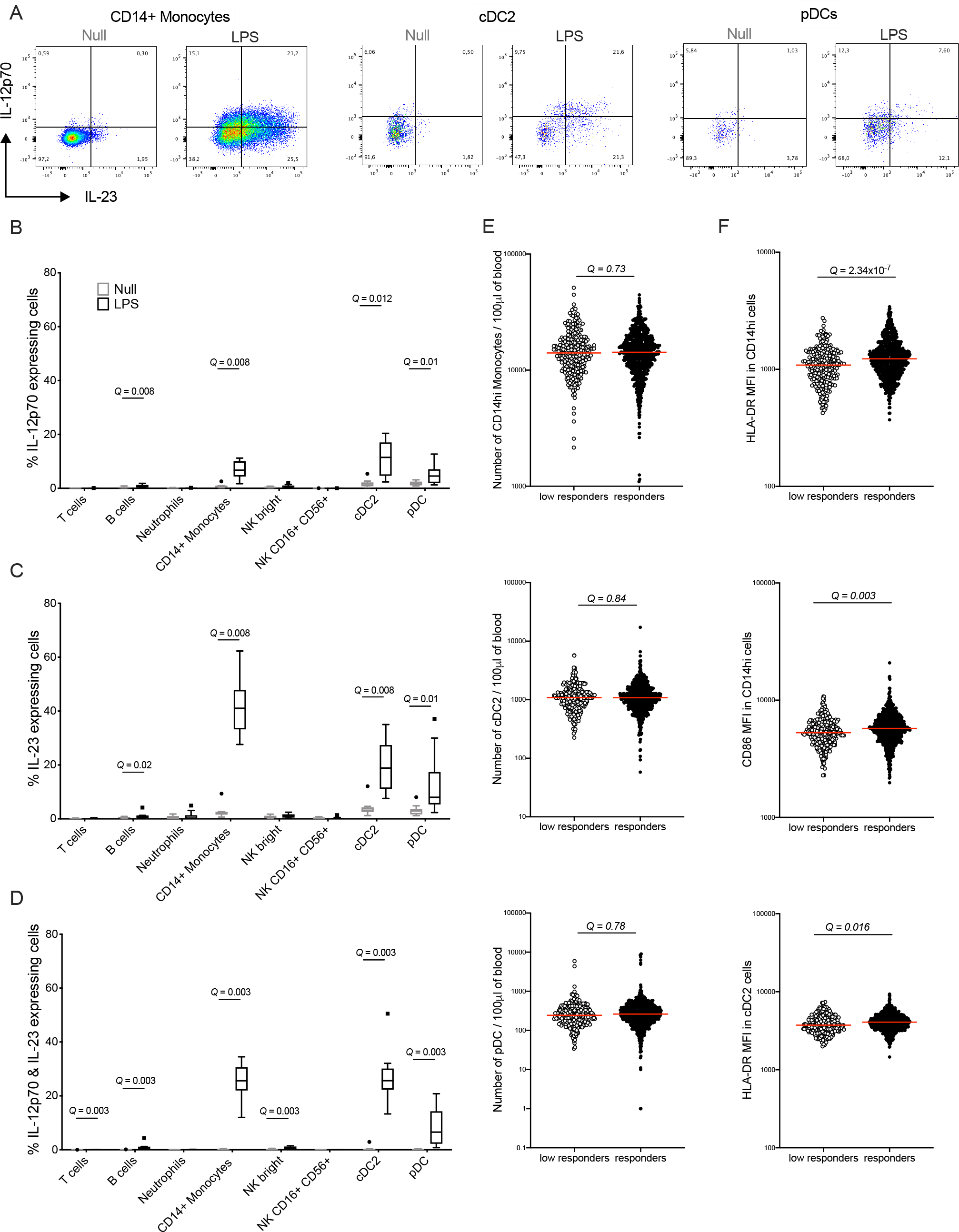
CD14^+^ monocytes and dendritic cells secrete IL-12p70 after LPS stimulation. See also Table S1. (**A**) Representative dot plots of IL-12p70 and IL-23 secretion upon null and LPS 22h stimulation measured by flow cytometry in CD14^+^ monocytes, cDC2, and pDCs. Tukey box-whisker plots of the percentage of IL-12p70^+^ cells (**B**), IL-23^+^ cells (**C**) or IL-12p70^+^ and IL-23^+^ cells (**D**) for the indicated immune cell populations in null and LPS conditions. Q values were determined by the Wilcoxon test and false discovery rate corrected for multiple comparison testing. (**E**) Numbers of CD14^hi^ monocytes, cDC2, and pDCs for the responder and low responder groups (n=1,000). (**F**) Expression levels of HLA-DR in CD14^+^ monocytes and cDC2 populations, and CD86 in CD14^+^ monocytes for the two groups. Red lines indicate the median value for each group. Q values were determined by an unpaired Student’s t test (on log10 values) and false discovery rate corrected for 79 (**E**) and 88 (**F**) multiple comparison testing (n=1,000).

Taking advantage of previously published data sets (Patin et al., 2018), we examined whether the low responders of the MI cohort had fewer circulating CD14^+^ monocytes or DCs than the responders at steady state. This immunophenotyping of the 1,000 donors was performed using ten flow cytometry panels from which 166 distinct immunophenotypes were reported (Patin et al., 2018). This included 76 absolute counts of circulating cells, 87 expression levels of cell surface markers and 3 cell ratios. No significant differences between the two groups were observed for numbers of CD14^+^ monocytes, cDC2 and plasmacytoid dendritic cells (pDCs) between low responders and responders (**Fig 2E**). However, an unbiased analysis applied to the entire immunophenotyping dataset, revealed significantly (Q < 0.05) lower cell surface expression of HLA-DR and CD86 on CD14^+^ monocytes and HLA-DR on cDC2 in the low responders as compared to the responders (**Fig 2F**).

Our analysis shows that CD14^+^ monocytes and DCs produce both IL-12p70 and IL-23 upon LPS stimulation. The numbers of these cells in the circulation of the low responders were not different, although they expressed less HLA-DR and CD86 activation markers.

### Five cis-acting single-nucleotide polymorphisms impact IL-12p70 protein secretion but not gene expression

Given the high prevalence of this phenotype, we next investigated for genetic associations utilizing existing genome-wide DNA data sets (Patin et al., 2018). Protein quantitative trait loci analysis (pQTL), including both cis and trans-located variants, was performed to find associations between IL-12p70 production as measured by Simoa after 22 hours of LPS stimulation and 5,265,361 genotyped single nucleotide polymorphisms (SNPs) (*P* value threshold of 4.32e-6 and 2.92e-9 for cis and trans effects, respectively). By mapping local cis-acting pQTLs (within 1 Mb of each gene coding for the *IL12A* and *IL12B* genes, which encode for the p35 and p40 subunits respectively), we found significant associations between five variable genetic regions, SNPs at rs17753641, rs17809756, rs17810546, rs76830965 and an indel at rs143060887 and IL-12p70 levels (*P* < 4.32e-6) (**Fig 3A**). Donors with the homozygous major allele for these 5 genetic regions had a significantly higher proportion (P < 0.0004) of IL-12p70 low responders compared to the heterozygous genotype group These five SNPs are all located near the *IL12A* gene and found to be in linkage disequilibrium in the human genome. We also performed a pQTL analysis using IL-12p70 concentrations measured in the 500 MI donors involved in the second visit and observed that these variants remained significantly associated with IL-12p70 production following LPS stimulation (**Fig S1A**). Importantly, none of these SNPs impacted IL-12p40 or IL-23 production (representative example of rs143060887 shown in **Fig 3B** and **Fig S1B**). We explored whether these genetic variants impacted the gene expression of the IL-12p70, IL-12p40, and IL-23 subunits. The expression of 560 immune-related genes was measured using Nanostring for the 1,000 MI donors after 22 hours of LPS stimulation. We found that the allelic dosage of the five SNPs did not impact the RNA expression levels of *IL12A, IL12B* and *IL23A* genes coding for p35, p40 and p19 subunits respectively (**Fig 3C** and **Fig S1C**).

**Figure 3.**
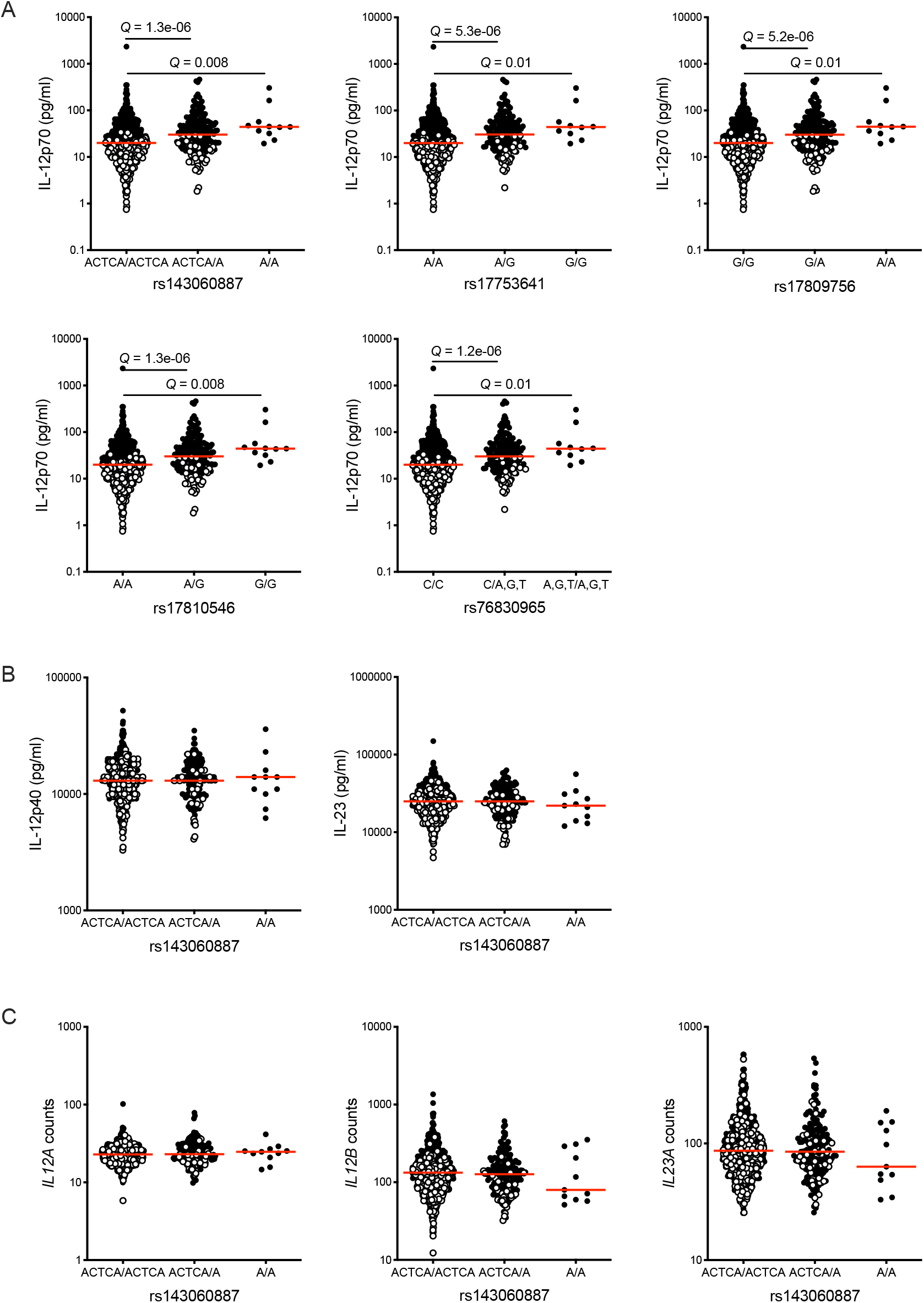
LPS-specific cis-acting pQTLs on IL-12p70 production. See also Figure S1. (**A**) Local pQTLs located near the *IL12A* gene acting specifically on IL-12p70 protein secretion measured by Simoa in the 1,000 *Milieu Interieur* donors in response to 22h LPS stimulation. (**B**) IL-12p40 and IL-23 protein measured by Luminex after 22h LPS stimulation according to the rs143060887 allelic dosage. (**C**) Dot plots show rs143060887 genotype-stratified gene expression levels for the *IL12A, IL12B, and IL23A* genes measured by Nanostring after 22h LPS stimulation. Empty circles represent IL-12p70 low responders of the *Milieu Interieur* cohort. Red lines indicate the median value for each group. Q values were determined by an unpaired Student’s t test using log10 values and false discovery rate correction was performed to correct for multiple comparison testing.

Together, our analysis indicated that the cis-acting variable region is associated with less IL-12p70 protein production but not gene expression after 22 hours of LPS stimulation in healthy donors.

### Variability in IL-12p70 is determined by upstream variability in IFNβ

To investigate potential molecular mechanisms behind variable IL-12p70 responses, we performed differential gene expression analysis between the responder and low responder groups. Nanostring analysis of the null condition revealed 52 differentially expressed genes (Q < 0.01) (**Table S2**). However, following LPS stimulation, 332 genes were significantly differentially expressed (**Fig 4A**) (**Table S3**). We found that gene expression of the p35, p40, and p19 subunits was significantly lower (*P* < 0.0001) in the low responders compared with the IL-12p70 responders after LPS 22 hours stimulation (**Fig S2A**) showing that variability in subunit transcripts may be responsible for the cytokine concentrations. IFNγ was the most differentially expressed gene with higher expression in the responders (Q = 7.34e-70 and fold change = 1.51). In addition, gene set enrichment analysis showed that 74 of the 332 differentially expressed genes were involved in the IFNγ response pathway (Q = 0.02) (**Fig S2B**).

**Figure 4.**
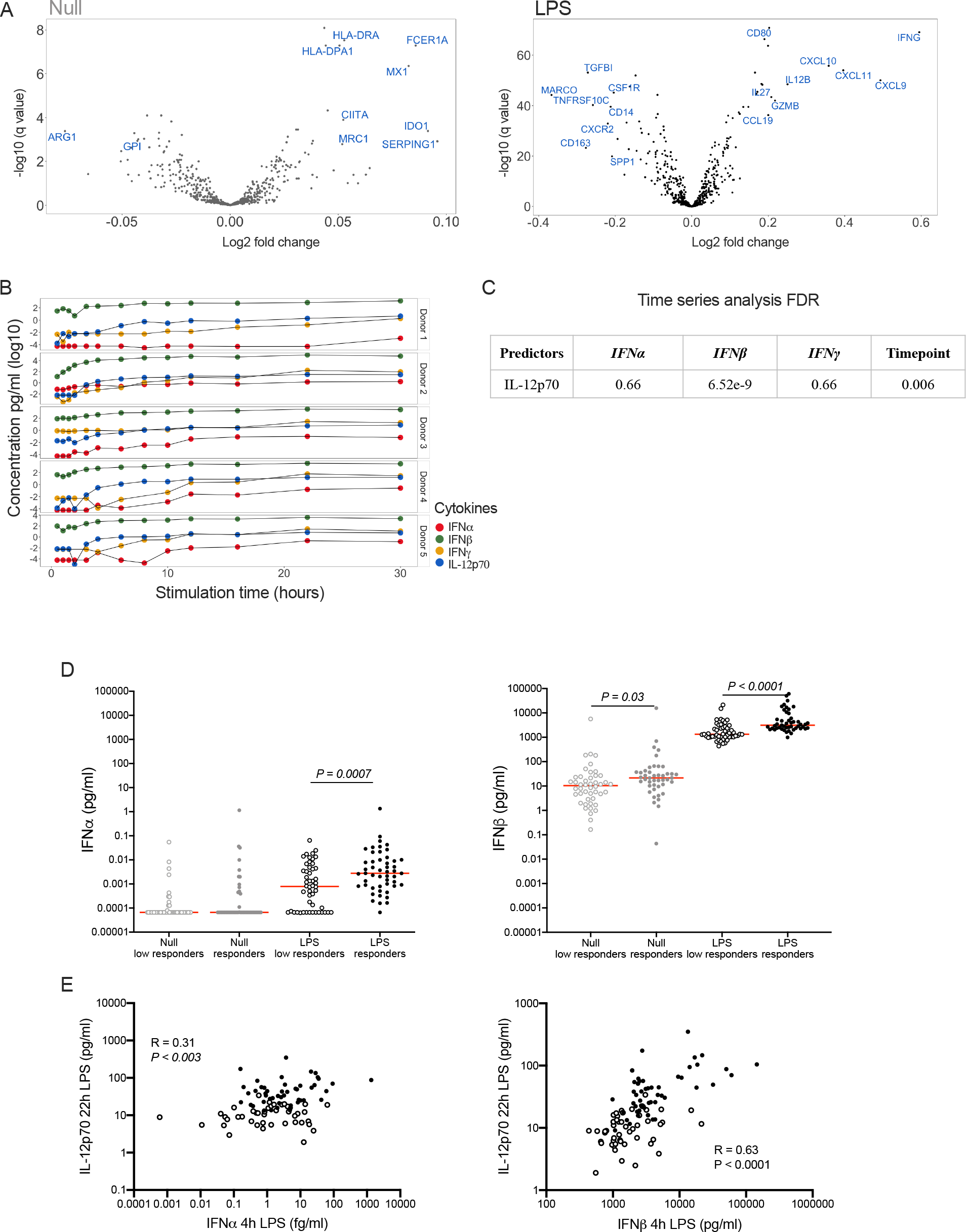
Kinetic responses and interactions of type I and II interferons and IL-12p70. See also Figure S2, Table S2 and Table S3. (**A**) Volcano plots displaying differentially expressed genes between *Milieu Interieur* IL-12p70 low responders and responders upon 22h null and LPS stimulation. Negative log2 fold change represents genes that are more expressed in low responders whereas positive log2 fold change represents genes that are more expressed in responders. The cut-offs used to display gene names are Q value < 0.01, log2 fold change > 0.05 in the null stimulation condition and log2 fold change > 0.2 in the LPS stimulation condition. (**B**) Type I (IFNα and IFNβ) and II (IFNγ) interferons and IL-12p70 protein secretion over time after LPS stimulation for 5 healthy donors. (**C**) Table reporting FDR Q values of time series analysis on protein secretion. (**D**) Type I IFNs measured by Simoa after 4h LPS stimulation of whole blood for the low responders and responders. The red lines indicate the median value for each group and stimulation condition. P values were determined by the unpaired Student’s t test, n=100 donors. (**E**) Correlation between IFNα or IFNβ secretion after 4 hours of LPS stimulation and IL-12p70 secretion upon 22h LPS stimulation. Pearson correlation tests were performed on log10 values; n=100 donors. Empty circles represent IL-12p70 low responders of the *Milieu Interieur* cohort.

Previous studies reported that a first wave of IFNγ is necessary to induce IL-12p70, which triggers IFNγ secretion, further inducing IL-12p70 through a positive feedback loop (Hayes et al., 1995) (Abdi et al., 2006) (Snijders et al., 1998). Moreover, type I IFN such as IFNα induces *IL12A* gene expression, which codes for the p35 subunit, in response to TLR activation (Gautier et al., 2005). To assess the roles of type I and II interferons in IL-12p70 secretion upon LPS stimulation, we performed a detailed kinetic analysis. Whole blood of 5 healthy donors was stimulated with LPS, and IL-12p70, IFNα, IFNβ, and IFNγ were measured at 14 timepoints from 0 to 30 hours using Simoa digital ELISA. In 4 out of 5 donors, both IFNγ and IFNα were produced either simultaneously or after IL-12p70, suggesting that these two cytokines are not responsible for IL-12p70 secretion. However, for all 5 donors, IFNβ secretion began 30 minutes to 1 hour before IL-12p70 induction (**Fig 4B**). These observations suggest a link between IFNβ and IL-12p70 secretion after LPS stimulation. To test this, we performed a time series analysis using a linear mixed-model approach which showed a significant association between IFNβ and IL-12p70 (Q = 6.52e-9), whereas no significant associations were found with either IFNγ or IFNα (Q = 0.66) (**Fig 4C**).

To determine if the low responders of the MI cohort secreted less type I IFNs, we measured IFNα and IFNβ proteins in null and LPS TruCulture supernatants after 4 hours of stimulation in a subset of 100 donors. Both IFNα and IFNβ protein were significantly lower in the low responder group (P = 0.0007 and P < 0.0001, respectively), although IFNβ was secreted to a much higher degree than IFNα (over 6 logs higher, and 160-fold induction for IFNβ compared to 20-fold for IFNα) (**Fig 4D**). Furthermore, early IFNβ production was positively and significantly correlated with later induction of IL-12p70 (Pearson’s R value 0.63, *P* < 0.0001) (**Fig 4E**). Together, our analyses indicate that low IFNβ production is associated with the IL-12p70 low responder phenotype in whole blood of healthy donors after LPS stimulation.

### The secretion of IFNβ by monocytes is essential for IL-12p70 secretion/Th1 response upon LPS stimulation

To test whether IFNβ was more essential than IFNα for LPS-induced IL-12p70 secretion, we performed additional whole blood stimulations. Addition of either IFNα or IFNβ to LPS stimulation resulted in a significant increase in IL-12p70 (Q = 0.0034 and Q < 0.0001 respectively), however the fold increase was a log (x9.8) greater with IFNβ (**Fig 5A**). Either cytokine alone did not induce IL-12p70, suggesting that type I IFNs can induce the production of only one of the IL-12p70 subunits. By contrast, the addition of IFNβ or IFNα to LPS stimulation did not increase IL-12p40 nor IL-23 secretion, suggesting that type I IFNs regulate production of the p35 subunit (**Fig S3A**). We further investigated the regulation of the p35 and p40 subunits by measuring their gene expression following 4 hours stimulation with IFNα and an unstimulated control (null) in 25 donors of the MI cohort. We observed that IFNα induced both *IL12A* and *IL12B* gene expression, but the effect size was greater for *IL12A* (Cohen’s *d* = 1.75 and 0.70 for *IL12A* and *IL12B* respectively) confirming that type I IFNs up-regulate the production of the p35 subunit (**Fig 5B**).

**Figure 5.**
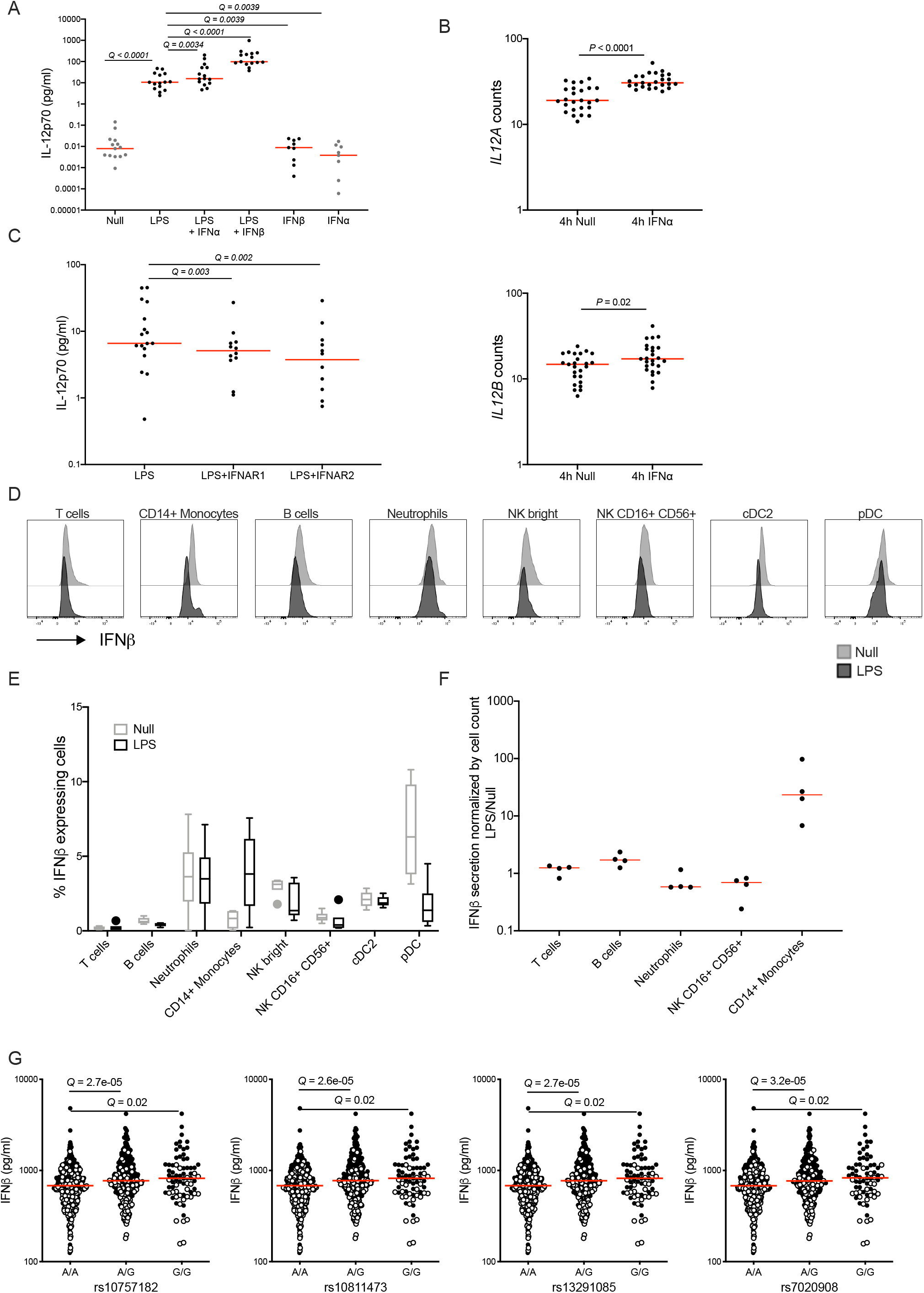
Impact of LPS, type I IFNs and IFNAR inhibitors on IL-12p70 production. See also Figure S3. (**A**) IL-12p70 secretion measured by Simoa in whole blood after 22h stimulation with LPS, type I IFNs, the combination of LPS and type I IFNs (n=9) and an unstimulated control (null). The red lines indicate the median value for each stimulation condition and Q values were determined using the Wilcoxon test and false discovery rate correction was performed to correct for multiple comparison testing. (**B**) *IL12A* and *IL12B* gene expression upon 4 hours whole-blood stimulation with IFNα and an unstimulated control (null), n=25. The red lines indicate the median value for each stimulation condition and P values were determined by the Wilcoxon test. (**C**) IL-12p70 secretion measured by Simoa in whole blood after 22 hours stimulation with LPS, LPS with IFNAR neutralizing antibodies, or an unstimulated control (null). The red lines indicate the median value for each stimulation condition and Q values were determined using the Wilcoxon test and false discovery rate correction was performed to correct for multiple comparison testing. (**D**) Representative histograms showing intracellular IFNβ production measured by flow cytometry in T cells, CD14^+^ monocytes, B cells, neutrophils, NK bright cells, NK cells, cDC2 and pDC cell populations after 6h of null or LPS whole blood stimulation. (**E**) Tukey box-whisker plots of the percentage of IFNβ cytokine positive cells for the indicated immune cell populations in null and LPS conditions, n=6. (**F**) Intracellular IFNβ (LPS/null ratio) measured by Simoa after 3h null and LPS whole blood stimulation, cell sorting, and lysis of purified cells, n=4 with a minimum of 100,000 purified cells per population. (**G**) Cis-acting variants significantly associated with LPS-induced IFNβ protein secretion measured in the 1,000 *Milieu Interieur* donors after 22h LPS stimulation.

To determine whether type I IFNs were essential for IL-12p70 production, we performed stimulation experiments while blocking the type I IFN pathway. Neutralization of the IFNAR1 or IFNAR2 subunits with either anti-IFNAR1 or anti-IFNAR2 mAbs (neutralizing capacity validated in **Fig S3B**), in the presence of LPS, significantly decreased IL-12p70 secretion (**Fig 5C**). This result confirmed that IFNβ signaling through IFN type I receptor is necessary for IL-12p70 secretion.

We next investigated which immune cells secrete IFNβ in whole blood of healthy donors after TLR4 activation by intracellular flow cytometry. For all donors, we observed IFNβ secretion by CD14^+^ monocytes upon LPS stimulation compared with the null condition, in which no IFNβ secretion was detected (**Fig 5D** and **5E**). To further confirm this result with a complementary approach, we purified 5 immune populations by flow cytometry based cell sorting after whole blood LPS stimulation and measured IFNβ in the cell lysates. Supporting our previous results, IFNβ secretion was only detected in CD14^+^ monocytes after LPS stimulation as compared to the unstimulated control (**Fig 5F**).

To provide insight into variability of the IFNβ response, we performed a cis- and trans-pQTL analysis after 4 hours of LPS stimulation in MI donors. We found significant associations between four genetic variants rs10757182, rs10811473, rs13291085 and rs7020908 and IFNβ levels (*P* < 4.32e-6) (**Fig 5G**). These SNPs are in linkage disequilibrium and are located within 1 Mb of the *IFNB1* gene coding for IFNβ. For all SNPs, no significant differences were observed regarding the proportion of IL-12p70 low responders among the 3 allelic dosages of each variant and none of the variants impacted *IFNB1* gene expression (**Fig S3C**).

We demonstrated that IFNβ production is crucial for induction of IL-12p70, and is mainly produced by monocytes after LPS stimulation of whole blood. In addition, the production of IFNβ protein is associated with a cis-acting variable genetic region.

### IKKb and IRF5 gene methylation impact the IL-12p70 phenotype

As epigenetic changes can drive cytokine secretion and control the level of immune responses (Morandini et al., 2016), we next assessed whether DNA methylation contributes to IL-12p70 secretion variability, utilizing gene methylation data sets from MI donors. For maximum statistical power, we focused our analysis on 49 selected genes involved in the LPS response pathway (listed in **Table S4**). Of these selected genes, we identified 29 probes (out of 1234 probes tested), corresponding to 20 unique genes (**Table S5**), that showed significantly different methylation levels between the IL-12p70 low responders and responders (Q < 0.05). Interestingly, 7 probes that showed increased methylation in the responder group were associated with *IKBKB* and *IRF5* genes (**Fig 6A**). The *IKBKB* gene codes for the IKKb kinase, which phosphorylates and activates the transcription factor IRF5 (Ren et al., 2014), which plays a role in inducing IFNβ expression (Bergstrom et al., 2015) (del Fresno et al., 2013). The methylation levels of these probes negatively correlated with *IKBKB* and *IRF5* gene expression in the LPS stimulation condition but not in the null control (example shown **Fig 6B**). This result highlights the impact of methylation on *IKBKB* and *IRF5* gene expression for downstream cytokine responses to LPS.

**Figure 6.**
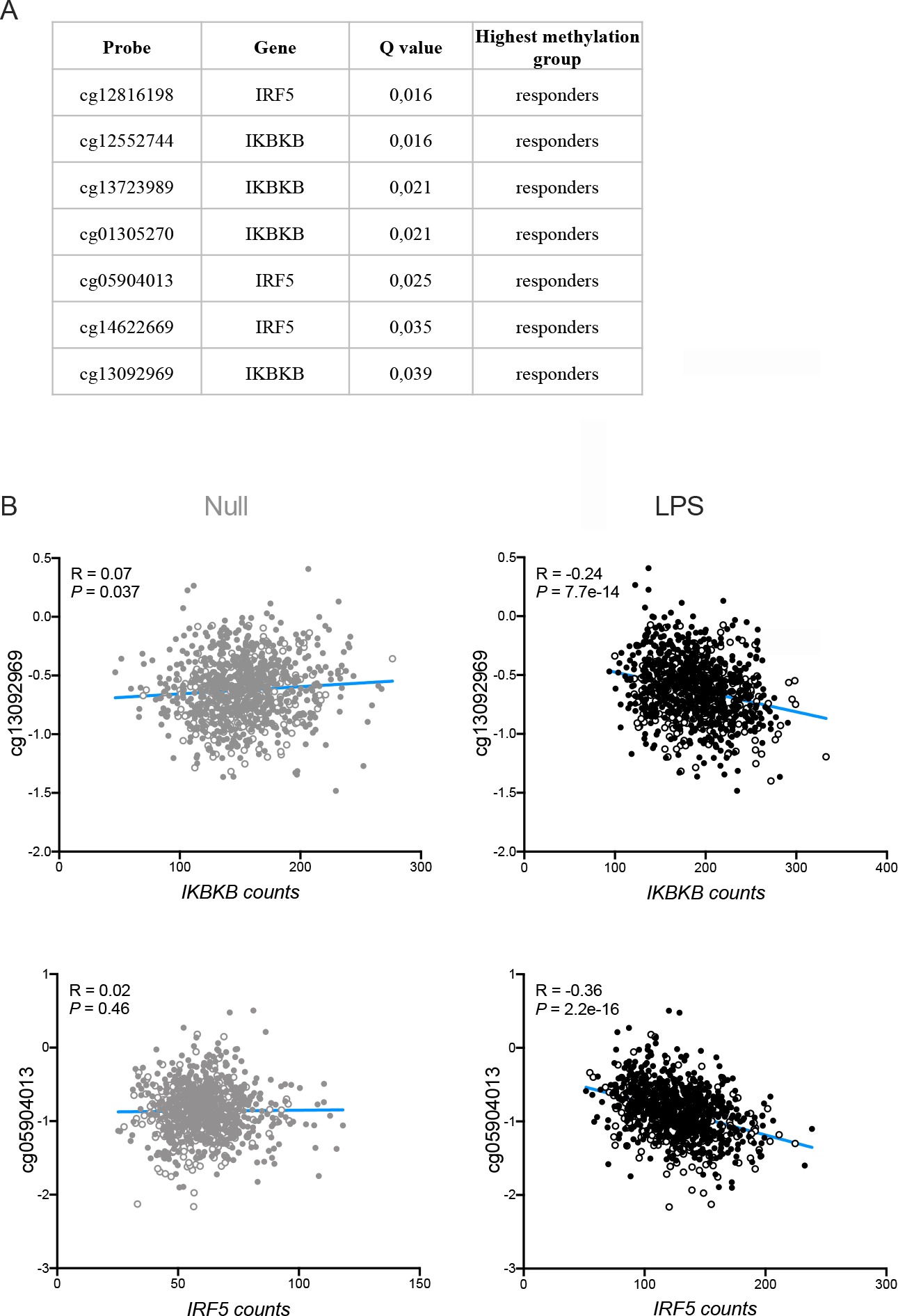
Differential methylation probes between low and high responders. See also Table S4 and Table S5. (**A**) List of *IKBKB* and *IRF5* gene methylation probes significantly different between the *Milieu Interieur* low responders and responders. (**B**) Correlation of one IKBKB and IRF5 probe methylation values with their respective gene expression measured by Nanostring after null and LPS 22h stimulations. Pearson correlations were performed using gene expression log10 values. Empty circles represent IL-12p70 low responders of the *Milieu Interieur* cohort.

### Integrative model to dissect respective contributions of variable IL-12p70 and IFNβ responses and validation of the IFNβ/IL-12p70 pathway in disease

To assess the relative importance of intrinsic and genetic factors identified in our different analyses, we utilized the feature selection algorithm Boruta (Kursa and Rudnicki, 2010). Based on previous studies highlighting the importance of age (Carr et al., 2016), sex (Giefing-Kroll et al., 2015), smoking (Qiu et al., 2017), and cytomegalovirus (CMV) status (Sylwester et al., 2005) for immune variability, these variables were also included in the model. While CMV status and smoking variables showed no importance for IFNβ or IL-12p70 cytokines, sex appeared to have importance for IFNβ but not IL-12p70 secretion (**Fig 7A**). IFNβ secretion, genetic variants, and HLA-DR MFI in monocytes were the top 3 variables showing high importance for variable IL-12p70 secretion, whereas HLA-DR MFI in monocytes and age were particularly important for IFNβ secretion. Selected features from the algorithm were integrated in a linear regression model to predict and explain the relationship between IFNβ or IL-12p70 secretion and the genetic and nongenetic independent variables (**Fig 7B**). Age, monocyte HLA-DR expression levels, and genetic variants showed the most significant associations with IFNβ secretion (P < 0.001). HLA-DR MFI in monocytes was also significantly linked with IL-12p70 production in addition to IFNβ and genetic variants (P < 0.001) confirming our previous analysis. Finally, we quantified the contribution of intrinsic and genetic factors to IFNβ and IL-12p70 response variation (**Fig 7C**). We found that the expression levels of cellular activation markers on monocytes and cDC2 had the strongest impact on IFNβ secretion, explaining 4.4% of the variance, whereas sex explained only 1.1% of the variance. The impact of IFNβ production was particularly strong for IL-12p70 secretion as it accounted for 21.6% of the total variance whereas genetics, epigenetics, and cellular markers accounted for less than 5% of the variance individually. In addition, age had a weak impact on IL-12p70 secretion. This analysis also revealed that 67% of variance in IL-12p70 and 88% of variance in IFNβ responses remains unexplained by these models.

**Figure 7.**
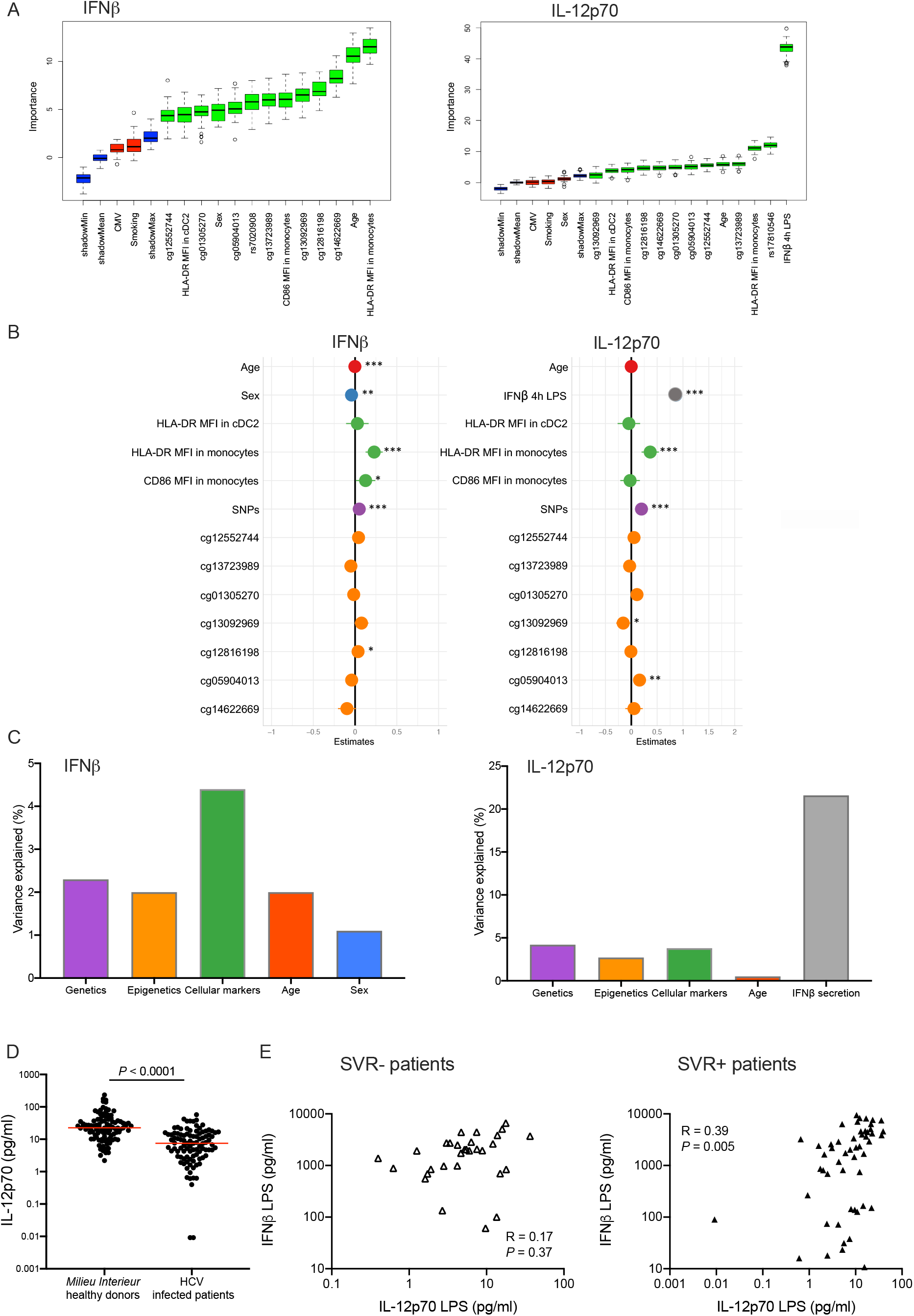
IFNβ secretion drives IL-12p70 variability both in healthy donors and HCV infection. See also Figure S4. (**A**) IFNβ and IL-12p70 Boruta variable importance charts, including all genetic and non-heritable factors selected from the exploratory analysis as well as age, sex, CMV status and smoking. Red and green boxplots represent Z scores of rejected and confirmed attributes respectively. Blue boxplots correspond to minimal, average and maximum Z scores of a shadow attribute. (**B**) Forest plots displaying the statistical significance of each variable associated with IFNβ and IL-12p70 resulting from the predictive linear regression models. (**C**) Proportion of the variance explained by genetic and non-genetic factors regarding IFNβ and IL-12p70 secretion following whole-blood LPS stimulation at 4h and 22h, respectively. (**D**) IL-12p70 measured using Simoa after stimulation of whole blood with LPS for 22h from healthy donors (n=100) and patients infected with HCV (n=100). P values were determined by Mann-Whitney tests. (**E**) Correlation between IFNβ and IL-12p70 secretion upon LPS stimulation in SVR-HCV patients (n=33 patients) and in SVR+ HCV patients (n=54 donors). Pearson correlation tests were performed on log10 values.

To determine whether the IL-12p70 response is also associated with IFNβ secretion in disease settings, we examined secretion of type I IFNs and IL-12p70 in HCV patients. This disease was selected as a type I interferon gene signature is involved in immunopathogenesis (Sarasin-Filipowicz et al., 2008). Furthermore, the HCV patients were treated with 6 months of pegylated interferon plus ribavirin, which was the standard of care treatment at the time of the clinical study. This provided the opportunity to test the applicability of our findings to a relevant therapeutic setting.

The patient cohort was established as described in the Methods and whole blood was stimulated for 22 hours with LPS and an unstimulated control (null) using the same TruCulture system as applied to MI donors, after which we measured IFNα, IFNβ, and IL-12p70 responses by Simoa. IL-12p70 cytokine secretion measured with the Simoa assay, was significantly lower (*P* < 0.0001) in patients infected with HCV than in healthy donors following LPS stimulation, suggesting that the Th1 response is altered in this disease (**Fig 7D**). Patients were divided into those that were sustained virological responders (SVR+) or not (SVR-) after 6 months of treatment with pegylated interferon plus ribavirin. No significant differences were observed in IL-12p70, IFNβ, or IFNα secretion between the SVR+ and SVR-patients for any of the stimulation conditions (**Fig S4A**). However, IFNβ positively and significantly correlated with IL-12p70 secretion after LPS stimulation in SVR+ patients (Pearson’s R value 0.39, *P* = 0.005) but not in the SVR-patients (Pearson’s R value 0.17, *P* = 0.37) (**Fig 7E**). By contrast, IFNα did not correlate with IL-12p70 secretion in SVR+ patients (Pearson’s R value 0.23, *P* = 0.1) but significantly correlated with IL-12p70 secretion in SVR-patients (Pearson’s R value 0.43, *P* = 0.01) (**Fig S4B**). These results show that the IFNβ-IL-12p70 pathway is altered in a subgroup of HCV infected patients who did not respond clinically to IFN-based treatment.

## Discussion

In this study, we dissected natural variability in IL-12p70 responses by applying a systems immunology approach to a population-based study. We characterized a consistently low IL-12p70 response upon LPS whole blood stimulation in a quarter of a healthy population cohort which allowed us to classify subjects into two groups regarding their immune responses to TLR4 activation. We also demonstrated that specific IFNβ signaling is key for triggering immune responses to bacteria, consistent with previous studies (Gautier et al., 2005) highlighting that type I IFNs are necessary for optimal IL-12p70 secretion.

We found that variable IFNβ secretion by monocytes in whole blood was the main driver of inter individual variability in IL-12p70 secretion following TLR4 activation, accounting for 21.6% of the total variance. This finding highlights the importance of integrating upstream cytokine networks for understanding variability in immune responses. The effects of genetics and epigenetics accounted for less than 10% of the total variance for both cytokines. Although this represents an important contribution, it does not support the hypothesis that genetic factors are the main determinant influencing cytokine responses, as previously reported (Li et al., 2016b). Here, we found that levels of monocyte HLA-DR and CD86 cell surface expression on circulating immune cells had a higher impact on induced IFNβ response variability than the combined effects of genetic and epigenetic factors. Low expression of HLA-DR and CD86 have been previously associated with impaired cellular functions of monocytes, including reduced pro-inflammatory properties and a decreased ability to induce T cell responses through antigen presentation (Piani et al., 2000) (Astiz et al., 1996). Thus, our results suggest that low responders may have an increased incidence or be at greater risk of bacterial infection. The importance of the IL-12p70-IFNγ axis in bacterial infection has been clearly demonstrated by previous studies of Mendelian susceptibility to mycobacterial infection (MSDM) (Martinez-Barricarte et al., 2018). As clinical examples of MSDM are extremely rare, our study shows how such information can be obtained at the population level to identify lower impact, but higher frequency, genetic variants that likely combine with other factors for an overall increased disease risk.

Highlighting the novelty of our approach, we report two independent genetic associations with both IL-12p70 and IFNβ protein secretion specific to LPS stimulation. All pQTLs described are cis-QTLs, suggesting that these pQTLs directly influence cytokine secretion, contrary to recent reports which identified trans-QTLs as having a stronger indirect influence on cytokine secretion (Li et al., 2016b) (Li et al., 2016a). We (Quach et al., 2016) and others (Fairfax et al., 2014) have previously reported a SNP (rs12553564) that associates with variable IFNβ gene expression in LPS-stimulated monocytes that has a downstream trans-effect on multiple immune genes. Surprisingly this SNP was not significantly associated with either IFNβ or IL-12p70 secretion in our study. This difference may be explained by the measurement of secreted proteins rather than RNA, or by the multi-cellular composition of blood analyzed, but clarifying this will require further studies.

The effects of age and sex on IFNβ and IL-12p70 production were relatively moderate. We observed that age had a higher impact on variance of both cytokines as compared to sex, suggesting that aging or environmental exposures may affect cytokine secretion more than sex. Variation in Th1 immune responses due to age has also been reported by Horst *et al*., who showed a significant decrease of IFNγ with age following whole blood LPS stimulation. Our results suggest that Th1 cytokine production is impacted by immunosenescence as previously documented (Rink et al., 1998) (Crooke et al., 2019).

Our study also presents some limitations. First, we cannot exclude the possibility that 28% of the healthy donors had been exposed to LPS prior to blood collection, leading to endotoxin tolerance (Kox et al., 2011, Rittig et al., 2015). Second, gene expression was determined for 560 immune genes, which does not provide an unbiased view of the transcriptome. In addition, the genotyping method used did not provide extensive information about rare genetic variants and structural variation. Thus, our systems immunology approach may be extended in future studies using RNA-sequencing and whole genome sequencing to address these limitations. Third, the dose of LPS used to stimulate whole blood (10 ng/ml) is higher than the dose that induces sepsis (Fullerton et al., 2016). Although selected by titration experiments to be in the biological response range (Duffy et al., 2014), this relatively high dose induces a strong cytokine response which may mask potential contributions of genetic and non-heritable determinants. As a consequence, the IFNβ/IL-12p70 observed variability may not fully reflect natural variability present during bacterial infection. Finally, a non-negligible part of IL-12p70 and IFNβ immune response variance remains unexplained by our models (∼67% and ∼88% respectively). Thus, additional investigations are required to assess the contribution of other determinants, more specifically the impact of environmental exposures, the effects of additional genetic control including unexplored transcripts and indels, and alternative epigenetic effects such as chromatin remodeling. Alternatively, we may also consider that such biological systems are inherently noisy, and that despite best efforts to standardize assays and integrate multiple levels of variability, we may not be able to explain and account for all variability (Eling et al., 2019). However, the application of our findings to HCV patients show that even an uncomplete understanding of variable immunity may be useful for better understanding in disease.

Overall, our study provides new findings on how IL-12p70 is produced and regulated against bacterial stimulation both in health and disease settings, and a better understanding of how intrinsic and genetic factors drive inter individual immune responses. The characterization of the determinants of cytokine variation in healthy subjects is essential to identify causes that lead to aberrant cytokine secretion in immune-related disorders for development of new treatments. The systems immunology model described here allows the dissection of such complex pathways in a population setting for direct relevance to human studies and clinical applications.

## Data Availability

The genotype data reported in this paper have been deposited in the European Genome-Phenome Archive (EGA; accession no. EGAS00001002460).

https://ega-archive.org/

## Acknowledgements

This study was funded with support from the French Government’s Investissement d’Avenir Program, Laboratoire d’Excellence “Milieu Intérieur” Grant ANR-10-LABX-69-01 and by an Agence National de Recherche foundation grant (CE17001002). We thank the UTechS CB of the Center for Translational Research, Institut Pasteur for supporting data generation, Pierre-Henri Commere for help with flow cytometry sorting, Aurelie Bisiaux for flow cytometry advice, and Dr Molly Ingersoll for scientific advice and critical reading of the manuscript. DD thanks Immunoqure for provision of the mAbs under an MTA for the Simoa IFN-α assay.

## Author contributions

CP designed and performed experiments, analysed and interpreted data and wrote the manuscript. AL performed specific experiments, analysis and interpreted data. BC, JB, VB, NS performed experiments. VR, JB, VSA, MK and EP analysed data. TJS, EM, and SP provided patient samples and analyzed data from clinical studies. MLA, LQM and DD conceived the whole study, obtained funding and provided guidance. DD designed and supervised the whole study, designed experiments, interpreted data and wrote the manuscript. All authors contributed to manuscript revision, read and approved the submitted version.

Author: The Milieu Intérieur Consortium†.

† The Milieu Intérieur Consortium¶ is composed of the following team leaders: Laurent Abel (Hôpital Necker), Andres Alcover, Hugues Aschard, Philippe Bousso, Nollaig Bourke (Trinity College Dublin), Petter Brodin (Karolinska Institutet), Pierre Bruhns, Nadine Cerf-Bensussan (INSERM UMR 1163 – Institut Imagine), Ana Cumano, Caroline Demangel, Christophe d’Enfert, Ludovic Deriano, Marie-Agnès Dillies, James Di Santo, Françoise Dromer, Gérard Eberl, Jost Enninga, Jacques Fellay (EPFL, Lausanne), Ivo Gomperts-Boneca, Milena Hasan, Magnus Fontes (Institut Roche), Gunilla Karlsson Hedestam (Karolinska Institutet), Serge Hercberg (Université Paris 13), Molly Ingersoll, Rose Anne Kenny (Trinity College Dublin), Olivier Lantz (Institut Curie), Frédérique Michel, Hugo Mouquet, Cliona O’Farrelly (Trinity College Dublin), Etienne Patin, Sandra Pellegrini, Stanislas Pol (Hôpital Côchin), Antonio Rausell (INSERM UMR 1163 – Institut Imagine), Frédéric Rieux-Laucat (INSERM UMR 1163 – Institut Imagine), Lars Rogge, Anavaj Sakuntabhai, Olivier Schwartz, Benno Schwikowski, Spencer Shorte, Frédéric Tangy, Antoine Toubert (Hôpital Saint-Louis), Mathilde Touvier (Université Paris 13), Marie-Noëlle Ungeheuer, Christophe Zimmer, Matthew L. Albert (In Sitro)§, Darragh Duffy§, Lluis Quintana-Murci§,

¶unless otherwise indicated, partners are located at Institut Pasteur, Paris

§co-coordinators of the Milieu Intérieur Consortium

Additional information can be found at http://www.milieuinterieur.fr/en

## Materials and methods

### The Milieu Interieur cohort

The 1,000 healthy donors of the *Milieu Interieur* cohort were recruited by BioTrial (Rennes, France) from September 2012 to July 2013. It is composed of 500 women and 500 men, including 200 individuals per decade of life, between 20 and 69 years of age. The donors were selected based on inclusion and exclusion criteria detailed elsewhere (Thomas et al., 2015). The recruitment was restricted to individuals from metropolitan French origin for three generations. Donors were defined as healthy according to medical history, clinical examination, laboratory results, and electrocardiography. General information about socio-demographic, health-related life habits, childhood disease, vaccination history and family health history were reported in an electronic case report form (eCRF). Half of the subjects were randomly selected (stratified by age and sex) to return for a visit 2 that took place between 2 to 6 weeks after visit 1. The clinical study was approved by the Comité de Protection des Personnes - Ouest 6 on June 13, 2012, and by the French Agence Nationale de Sécurité du Médicament on June 22, 2012, and was performed in accordance with the Declaration of Helsinki. The study was sponsored by the Institut Pasteur (Pasteur ID-RCB Number 2012-A00238-35) and conducted as a single-center study without any investigational product. The original protocol is registered under ClinicalTrials.gov (NCT01699893). Informed consent was obtained from the participants after the nature and possible consequences of the studies were explained. The samples and data used in this study were formally established as the *Milieu Interieur* biocollection (NCT03905993), with approvals by the Comité de Protection des Personnes – Sud Méditerranée and the Commission nationale de l’informatique et des libertés (CNIL) on April 11, 2018.

### Hepatitis C cohort

Patients infected with hepatitis C virus were recruited by four different hospitals/centers in Paris Ile-de-France, France. The cohort included 100 donors from 18 to 70 years old infected with genotype 1 or 4 of the virus. Blood of the hepatitis C infected patients was collected prior to interferon treatment initiation. Written informed consent was obtained from all study participants.

### Healthy human fresh whole blood

For experimental validation, fresh whole blood was collected into sodium heparin tubes from healthy French volunteers enrolled at the Clinical Investigation and Access to BioResources (ICAReB) platform (Center for Translational Research, Institut Pasteur, Paris, France). These donors were part of the CoSImmGEn cohort (NCT03925272). Written informed consent was obtained from all study participants.

### Immunophenotyping

Ten flow cytometry panels of eight-colors each were developed in order to enumerate and phenotype the major circulating leukocyte populations of the 1,000 *Milieu Interieur* donors. Premix of antibodies were manually made and staining protocol was performed using the Freedom Evo 150 liquid handling system (Tecan). Samples were acquired using MACSQuant analyzers. Panel antibodies and gating strategies were done as previously reported (Hasan et al., 2015). Converted FCS format files of 313 immunophenotypes were analyzed using FlowJo software version 9.5.3, from which a total of 166 flow cytometry measures were retained including 87 MFI, 76 cell counts and 3 cell ratios (Patin et al., 2018).

### Genotyping

DNA genotyping of 719,665 SNPs was performed using the HumanOmniExpress-24 BeadChip (Illumina) for the 1,000 *Milieu Interieur* donors. To improve coverage of rare SNPs, 966 of the 1,000 donors were also genotyped at 245,766 exonic SNPs using the HumanExome-12 BeadChip (Illumina). After quality control filters previously described, the final data set included 5,265,361 QC-filtered SNPs (Piasecka et al., 2018).

### Whole Genome DNA methylation profiling

Genomic DNA of the 1,000 *Milieu Interieur* donors was extracted and treated with sodium bisulfite (Zymo Research) and bisulfite-converted DNA was applied to the Infinium MethylationEPIC BeadChip (Illumina, California, USA), using the manufacturer’s standard conditions. The MethylationEPIC BeadChip measures single-CpG resolution DNAm levels at 866,836 CpG sites in the human genome. DNA methylation data were analyzed as described in Bergstedt *et al*. (manuscript in preparation).

### Whole blood TruCulture stimulation

TruCulture tubes (Myriad RBM) containing LPS-EB (ultrapure) (10ng/ml), heat killed E. coli 0111:B4 (10^7^ bacteria), and Poly I:C (20ug/ml) (all Invivogen) dissolved in 2ml of buffered media were batch produced as previously described. Tubes were thawed at room temperature and 1ml of fresh blood was distributed into each tube within 15 min of collection. Tubes were mixed by inverting them and incubated at 37°C (±1°C) for 4 or 22 hours in a dry block incubator. After the incubation time, a valve was manually inserted into the tube to separate the supernatant from the cells. Supernatant was collected, aliquoted and immediately stored at - 80°C for protein secretion analysis. Cell pellets of the TruCulture tubes were resuspended in 2ml of Trizol LS (Sigma) and tubes were vortexed for 2 min at 2000 rpm and stored at -80°C for gene expression analysis. The kinetic study of gene expression and cytokines was previously described (Bisiaux et al., 2017).

### Gene expression analysis

Total RNA was extracted from the null and LPS TruCulture cell pellets for the 1,000 *Milieu Interieur* donors using NucleoSpin 96 miRNA kit (Macherey-Nagel). RNA concentrations were measured using Quantifluor RNA system kit (Promega) and RNA integrity numbers were determined using the Agilent RNA 6000 Nano kit (Agilent Technologies). Total RNA samples were analyzed using the Human Immunology v2 panel profiling 594 immunology-related human genes (Nanostring). Gene expression data were normalized as previously described (Piasecka et al., 2018). To identify gene expression differences between responders and low responders, a multiple linear regression approach was performed where cell proportions from the lineage panel (leukocytes, B cells, T cells, NK cells, monocytes, neutrophils) were used to regressed out any gene expression differences due to cell population differences between donors. The analysis was implemented using the R package “broom”’ v0.5.2. Multiple testing correction was then applied to select the significant genes.

### Protein secretion analysis

Supernatants from whole-blood TruCulture supernatants were analyzed by Myriad RBM (Austin, Texas, US) using the Luminex xMAP technology. Samples were analyzed according to the Clinical Laboratory Improvement Amendments (CLIA) guidelines. The least detectable dose (LDD), lower limit of quantification (LLOQ) and lower assay limit (LAL) were defined as previously described (Duffy et al., 2014). To quantify interferons (α, β, γ), IL-12p70, IL-12p40 and IL-23 at ultrasensitive concentrations, Single Molecule array (Simoa) technology was used and hombrew assays developed as previously described and reported in Table S6. All the assays were run on a 2-step configuration on the Simoa HD-1 Analyzer (Quanterix, US). Limit of detection (LoD) was defined (using the highest bottom value of 95% CI in Prism 8).

### Protein secretion and gene expression kinetic analysis

Time series analysis on the protein secretion and gene expression measurements, across 5 different donors, was conducted using a linear mixed-model approach. The time dependency was modelled by incorporating the parameter time as a continuous linear predictor, alongside the other protein or gene predictors, and the donors were modelled as a random effect. The model was implemented using the R package « nlme » v3.1.140.

### Type I IFNs pathway inhibition

Whole blood was diluted 1:3 with RPMI in the presence or absence of IFNAR1 (Anifrolumab) and IFNAR2 (PBL, clone MMHAR-2) blocking antibodies at the final concentration of 10 µg/ml. IFNα, IFNβ and IFNγ were added 1 hour after the IFNAR1 and IFNAR2 antibodies at the final concentration of 1000 IU/ml. Samples were incubated at 37°C for 22 hours and supernatants were collected for protein secretion analysis.

### Intracellular cytokine staining

Fresh heparinized whole blood of healthy donors was diluted 1:3 with RPMI + GlutaMAX (Gibco, Life Technologies) without any stimulant (null) or with 10 ng/ml of LPS (InvivoGen). Diluted blood was incubated at 37°C and Brefeldin A (GolgiPlug, BD Biosciences) was added 30 min post stimulation to stain for IFNβ cytokine or 8 hours after the beginning of the stimulation to stain for IL-12p70 and IL-23 cytokines. After 6h or 22h of stimulation, to detect IFNβ or IL-12p70 and IL-23 cytokines respectively, whole blood was centrifuged at 500g for 5 min at room temperature and supernatant was removed. 2mM of EDTA (Life Technologies) was added and samples were incubated at 37°C for 10 min to detach adherent cells from the tubes. Red blood cells were lysed during 10 min using Pharm Lyse reagent (BD Biosciences). Leukocytes were then washed with PBS 1X (Gibco, Life Technologies). Live dead fixable green (Thermo Fisher Scientific) was added 1:1000 into the cell suspension and samples were incubated for 30 min at 4°C protected from light. Cells were washed once with fresh FACS buffer (PBS + 2% FBS + 0,2mM EDTA). FcR blocking reagent (Miltenyi) was added to the resuspended cells followed by 10 min incubation at 4°C in the dark. Surface antibody premix was distributed and cells were incubated in the dark for 30 min at 4°C. Thereafter, cells were washed with cold PBS and resuspended into fixation/permeabilization solution (Cytofix/Cytoperm kit, BD Biosciences). Samples were incubated at 4°C for 15 min protected from light. Cells were then washed with cold PBS and resuspended in 1X perm/wash buffer (Cytofix/Cytoperm kit, BD Biosciences). A second FcR blocking step was performed on resuspended cells for 10 min at 4°C in the dark prior to performing the intracellular staining. Antibodies specific to cytokines were added with 1X perm/wash buffer (100 µl total volume) and samples were incubated for 1h at 4°C in the dark. Samples were vortexed once half way through the incubation. Cells were washed once with cold PBS and resuspended in cold PBS and immediately acquired on the cytometer. Flow cytometry data were generated using LSR Fortessa (BD Biosciences) and fcs files were analyzed using FlowJo software version 10.4.2 (Hasan et al., 2015). Statistical graphs were done using Prism 8.

### pQTL analysis

pQTL analyses were performed using the MatrixEQTL R package (Shabalin, 2012), with detection thresholds of 1e-3 for cis-pQTLs located within 1 Mb of each gene and 1e-5 for trans-pQTLs and age, sex, smoking status and CMV serology status as covariates. For annotations Ensembl gene ids ENSG00000171855, ENSG00000168811 and ENSG00000113302 were used respectively for *IFNB1, IL12A* and *IL12B* genes. Protein expression data was log transformed prior to pQTL search. Bonferonni correction for multiple testing was applied to the results and pQTLs were considered significant if *P* < 4.32e-6 (FDR<0 .05) for cis-pQTLq and *P* < 2.92e-9 (FDR<0 .05) for trans-pQTLs.

### Integrative modelling

Variable selection was performed using the feature selection algorithm Boruta package of R (Kursa and Rudnicki, 2010). All selected variables were tested for multicollinearity by computing the variance inflation factor using the caret package in R. IFNβ and IL-12p70 selected variables were included in a linear regression model. To confirm that the model was appropriate for the data, patterns of residuals diagnostic plots were checked. Finally, the proportion of the variance explained by each category of variable was determined by averaging the sums of squares in all orderings of the variables in the linear model using the lmg metric in the relaimpo package of R (Gromping, 2006).

## Supplemental information

**Figure S1, related to Figure 3.**
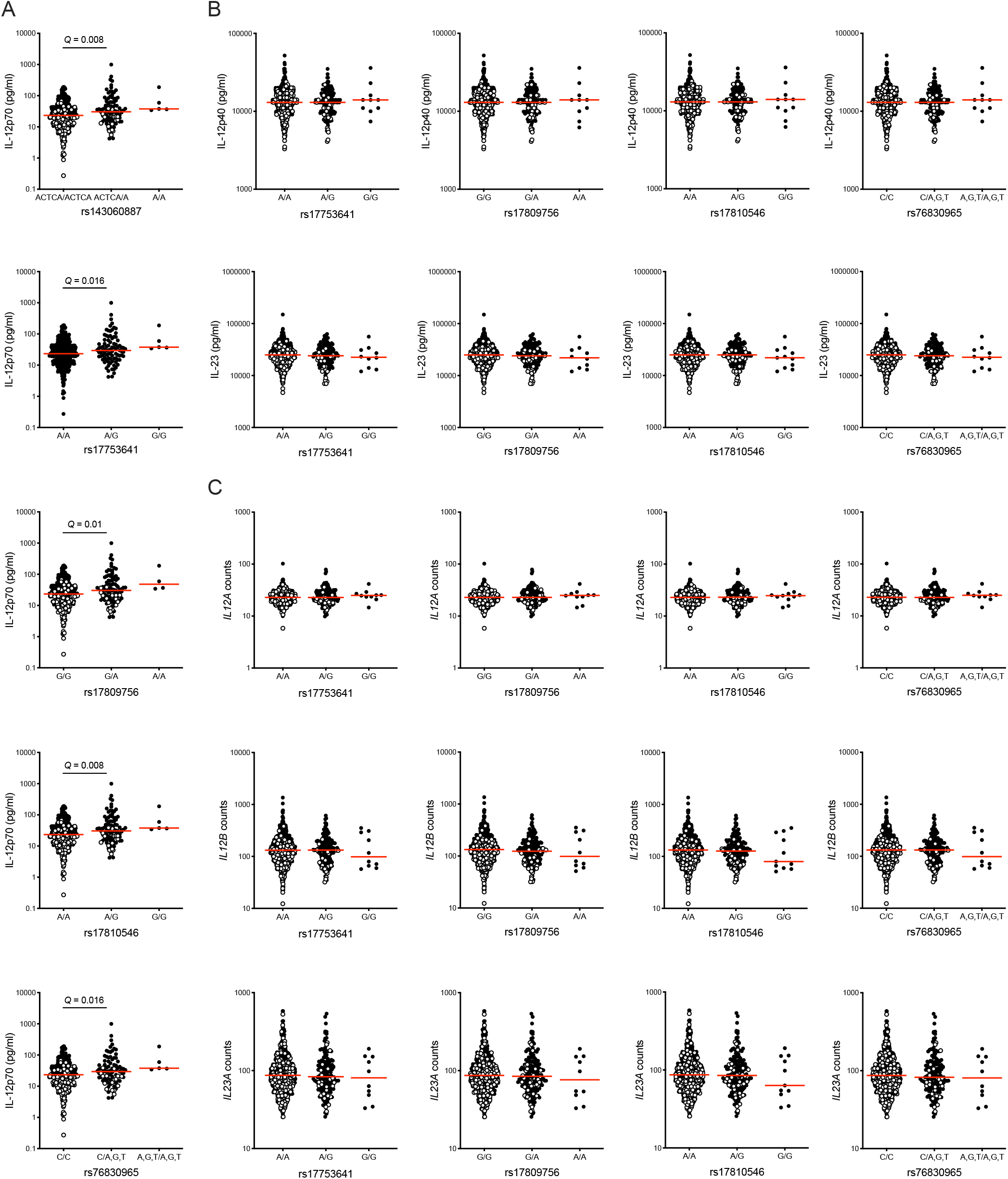
LPS-specific cis-acting pQTLs on IL-12p70, IL-12p40 and IL-23 production. (**A**) Local pQTLs located near the *IL12A* gene acting specifically on IL-12p70 protein secretion measured in the 500 *Milieu Interieur* donors who were sampled at a second time point in response to 22h LPS stimulation. (**B**) IL-12p40 and IL-23 protein secretion in the *Milieu Interieur* individuals following 22h LPS stimulation according to the rs17753641, rs17809756, rs17810546 and rs76830965 genotypes. (**C**) *IL12A, IL12B, and IL23A* gene expression measured by Nanostring after 22h LPS stimulation according to the rs17753641, rs17809756, rs17810546 and rs76830965 genotypes. Empty circles represent IL-12p70 low responders of the *Milieu Interieur* cohort. Red lines indicate the median value for each group. Q values were determined by an unpaired Student’s t test (on log10 values) and false discovery rate correction was performed to correct for multiple comparison testing.

**Figure S2, related to Figure 4.**
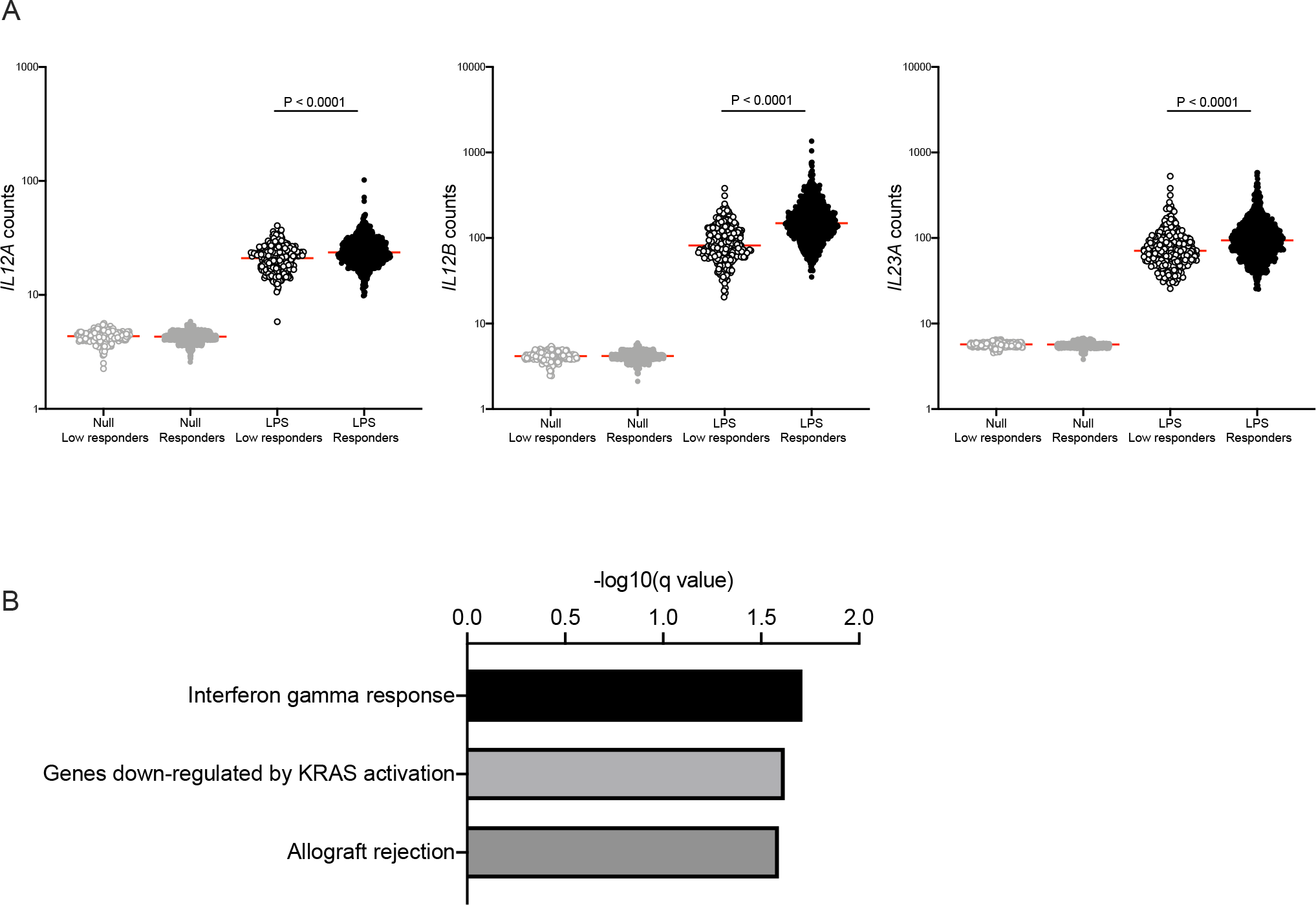
Differential gene expression analysis between the IL-12p70 low and high responders. (**A**) *IL12A, IL12B* and *IL23A* gene expression measured by Nanostring in the two groups of *Milieu Interieur* IL-12p70 responders after null and LPS stimulation. Red lines indicate the median value for each group. P values were determined by an unpaired Student’s t test on log10 values. (**B**) Gene set enrichment analysis bar plot displaying the significant pathways from the comparative analysis between the low responders and the responders following LPS 22h stimulation using the Molecular Signatures Database Hallmark gene sets.

**Figure S3, related to Figure 5.**
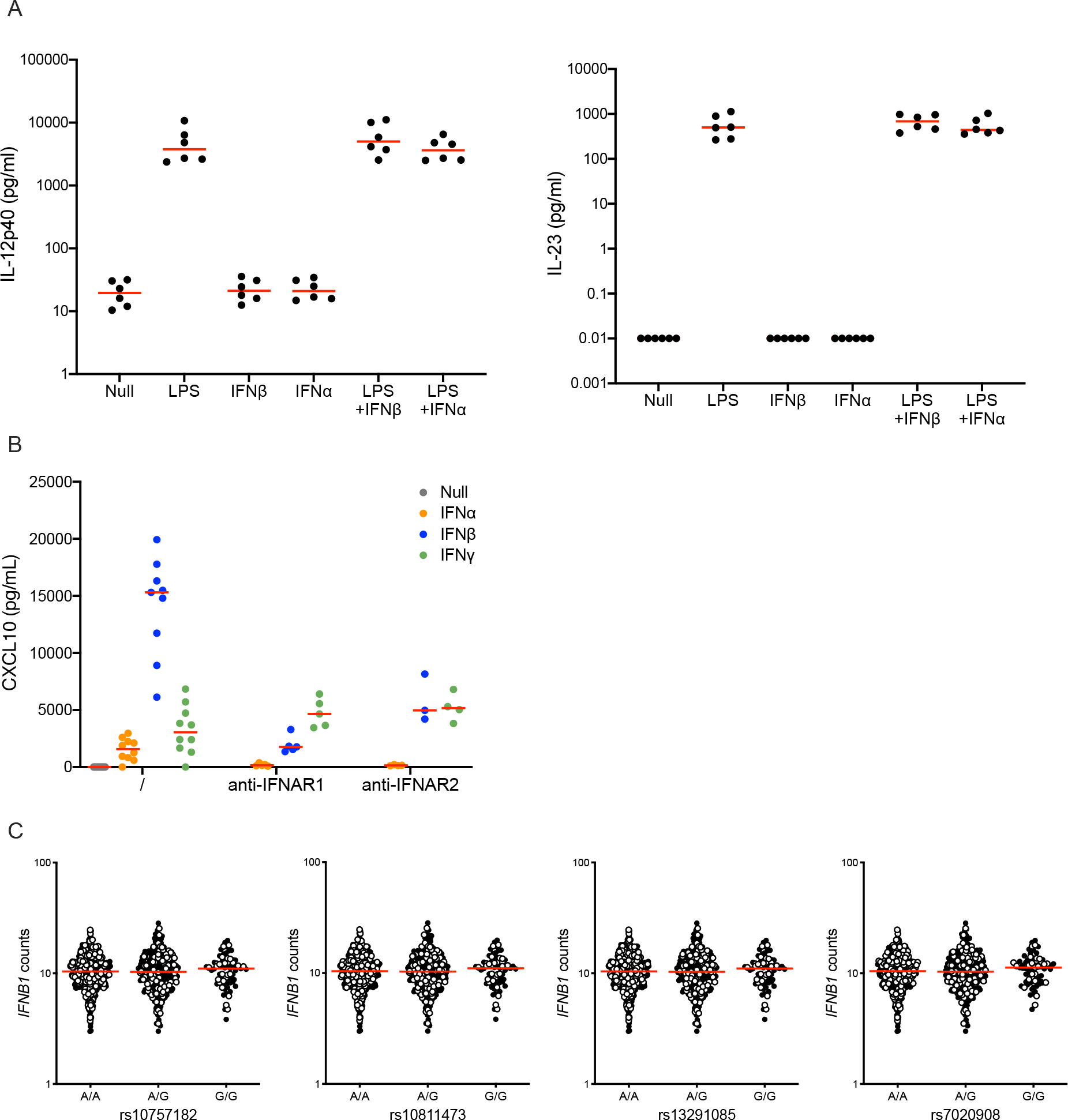
Impact of LPS and type I IFNs on IL-12p40 and IL-23 secretion. (**A**) IL-12p40 and IL-23 protein secretion measured by Simoa in whole blood after 22h stimulation with LPS, type I IFNs, the combination of LPS and type I IFN and an unstimulated control (null), n=6. The red lines indicate the median value for each stimulation condition. (**B**) CXCL10 protein secretion measured by Simoa in whole blood after 22h stimulation with type I and II IFNs in the presence or absence of IFNAR1 and IFNAR2 blocking antibodies. The red lines indicate the median value for each stimulation condition. (**C**) Dot plots show rs10757182, rs10811473, rs13291085 and rs7020908 genotype-stratified gene expression levels for the *IFNB1* gene measured by Nanostring after 22h LPS stimulation. Empty circles represent IL-12p70 low responders of the *Milieu Interieur* cohort. Red lines indicate the median value for each group. Q values were determined by an unpaired Student’s t test (on log10 values) and false discovery rate correction was performed to correct for multiple comparison testing.

**Figure S4, related to Figure 7.**
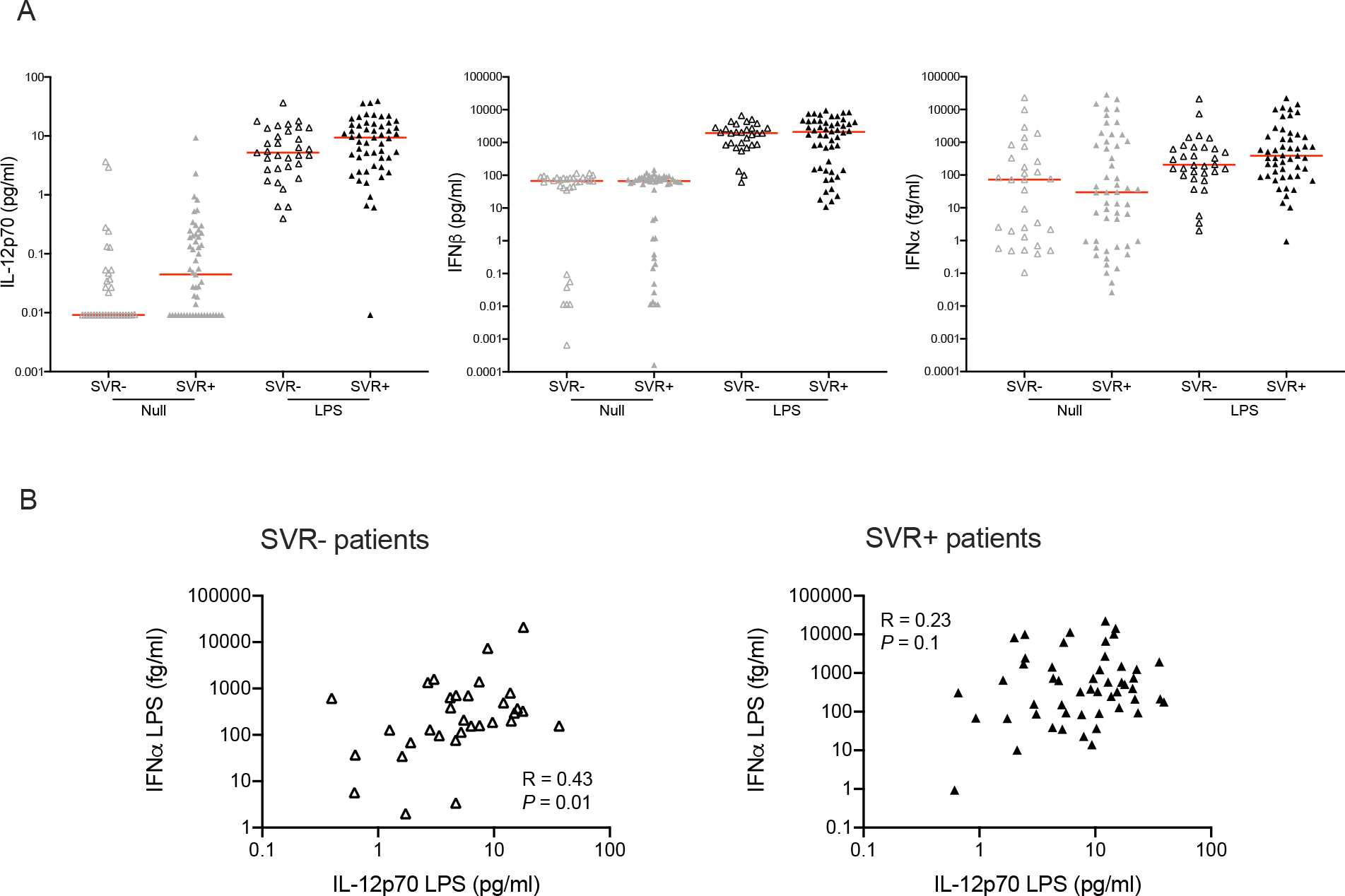
Type I IFNs and IL-12p70 responses in HCV infection. (**A**) IL-12p70, IFNβ and IFNα secretion measured by Simoa after 22h null and LPS stimulation of whole blood in SVR+ (n=54) and SVR-(n=33) HCV infected patients defined after 6 months of IFNα-based therapy. The red lines indicate the median value for each stimulation and group condition. P values were determined by the unpaired t test. (**B**) Correlation between IFNα and IL-12p70 secretion upon LPS stimulation in SVR-HCV patients (n=33 patients) and in SVR+ HCV patients (n=54 donors). Pearson correlation tests were performed on log10 values.

**Table S1, related to Figure 2.** IL-12p70 and IL-23 intracellular staining panel.

| **Marker** | **Color** | **Clone** | **Company** | **Dilution used**<br>**/100µl of blood** |
| --- | --- | --- | --- | --- |
| CD3 | APC-H7 | SK7 | BD Biosciences | 1/40 |
| CD19 | BV711 | SJ25C1 | BD Biosciences | 1/40 |
| CD56 | Pe-Cy7 | B159 | BD Biosciences | 1/20 |
| CD14 | V500 | M5E2 | BD Biosciences | 1/20 |
| CD16 | PE-CF594 | 3G8 | BD Biosciences | 1/40 |
| CD66b | BV421 | G10F5 | BD Biosciences | 1/50 |
| HLA-DR | PerCP-Cy5.5 | L243 | BD Biosciences | 1/10 |
| BDCA1 (CD1c) | BUV395 | F10/21A3 | BD Biosciences | 1/20 |
| BDCA2 (CD303) | AF700 | 201A | BioLegend | 1/40 |
| BDCA4 (CD304) | BV605 | U21-1283 | BD Biosciences | 1/20 |
| IL-12p70 | APC | REA123 | Miltenyi | 1/40 |
| IL-23p19 | PE | 23dcdp | eBioscience | 1/40 |

**Table S2, related to Figure 4.** Genes differentially expressed between the *Milieu Interieur* low responders and responders following whole blood Null stimulation.

**Table S1, related to Figure 2.** IL-12p70 and IL-23 intracellular staining panel.
| Gene | Log2 fold change | Q value |
| --- | --- | --- |
| HLA-DMB | 0,044 | 8,07E-09 |
| HLA-DRA | 0,053 | 2,96E-08 |
| FCER1A | 0,086 | 5,12E-08 |
| HLA-DPB1 | 0,044 | 5,12E-08 |
| HLA-DPA1 | 0,051 | 5,12E-08 |
| MX1 | 0,083 | 4,40E-07 |
| HLA-DRB3 | 0,045 | 4,73E-05 |
| IGF2R | -0,038 | 8,02E-05 |
| CEBPB | -0,032 | 8,02E-05 |
| CIITA | 0,052 | 0,0001 |
| ITGA5 | -0,027 | 0,0002 |
| SLAMF6 | 0,031 | 0,0004 |
| IFITM1 | 0,031 | 0,0004 |
| IDO1 | 0,092 | 0,0004 |
| ARG1 | -0,076 | 0,0004 |
| IL6R | -0,033 | 0,0005 |
| GAPDH | -0,044 | 0,0005 |
| MAPKAPK2 | -0,022 | 0,0006 |
| HLA-DMA | 0,027 | 0,0007 |
| NFIL3 | -0,033 | 0,0009 |
| IL1R1 | -0,040 | 0,0009 |
| S100A8 | -0,042 | 0,0010 |
| IFIH1 | 0,038 | 0,0012 |
| TNFRSF10C | -0,043 | 0,0012 |
| IFNGR1 | -0,028 | 0,0012 |
| SERPING1 | 0,096 | 0,0012 |
| LAIR1 | 0,026 | 0,0013 |
| CD74 | 0,038 | 0,0015 |
| MRC1 | 0,052 | 0,0016 |
| TNFRSF1B | -0,033 | 0,0020 |
| STAT6 | -0,029 | 0,0022 |
| S100A9 | -0,037 | 0,0023 |
| FCAR | -0,046 | 0,0024 |
| IRF5 | 0,024 | 0,0032 |
| LTB4R | -0,030 | 0,0034 |
| GPI | -0,050 | 0,0034 |
| CD82 | -0,031 | 0,0038 |
| CD83 | 0,035 | 0,0040 |
| IFI35 | 0,036 | 0,0048 |
| LILRB2 | -0,023 | 0,0048 |
| TYK2 | -0,022 | 0,0053 |
| CD81 | 0,021 | 0,0061 |
| IRF8 | 0,023 | 0,0070 |
| CD4 | 0,023 | 0,0082 |
| PRKCD | -0,025 | 0,0085 |
| BTLA | 0,023 | 0,0085 |
| LITAF | -0,023 | 0,0085 |
| TNFSF4 | -0,024 | 0,0085 |
| MME | -0,047 | 0,0085 |
| BST2 | 0,022 | 0,0086 |
| TAGAP | 0,023 | 0,0092 |
| RAF1 | -0,024 | 0,0095 |

**Table S3, related to Figure 4.**
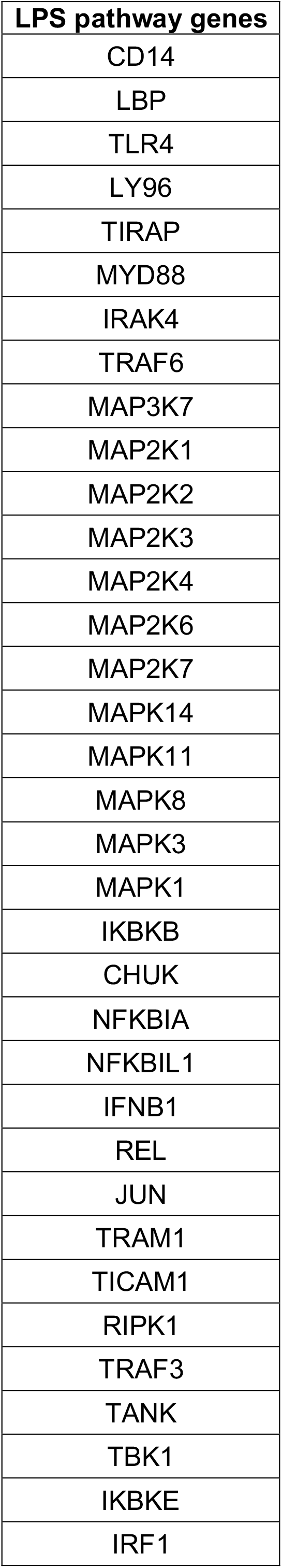
Genes differentially expressed between the *Milieu Interieur* low responders and responders following whole blood LPS stimulation.

**Table S4, related to Figure 6.**
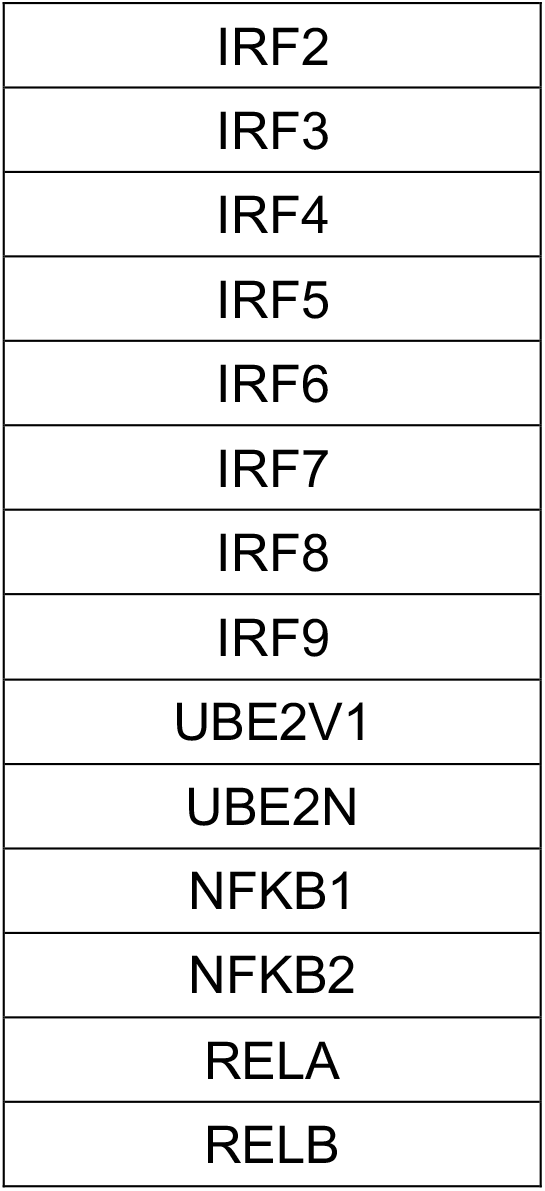
List of genes implicated in the LPS response pathway used to perform DNA methylation differential analysis between the low responders and the responders.

**Table S5, related to Figure 6.** Probes differentially methylated between the low responders and the responders.

| Probe | Gene | Q value |
| --- | --- | --- |
| cg12816198 | IRF5 | 0,016 |
| cg12552744 | IKBKB | 0,016 |
| cg21045241 | RELB | 0,021 |
| cg01037764 | TRAM1 | 0,021 |
| cg13723989 | IKBKB | 0,021 |
| cg01305270 | IKBKB | 0,021 |
| cg21053741 | NFKBIL1 | 0,021 |
| cg16604658 | TBK1 | 0,025 |
| cg04666274 | MAP2K2 | 0,025 |
| cg05904013 | IRF5 | 0,025 |
| cg06392169 | IRF4 | 0,025 |
| cg19308985 | IRF8 | 0,035 |
| cg20567119 | TRAF6 | 0,035 |
| cg16289653 | MAP2K2 | 0,035 |
| cg11917175 | IRF2 | 0,035 |
| cg14622669 | IRF5 | 0,035 |
| cg14456745 | TANK | 0,035 |
| cg16145324 | MAPK14 | 0,035 |
| cg24064264 | IRF7 | 0,035 |
| cg13730105 | TLR4 | 0,037 |
| cg10080401 | MAPK1 | 0,037 |
| cg04639174 | TRAF6 | 0,037 |
| cg13092969 | IKBKB | 0,039 |
| cg04123016 | MAP2K1 | 0,041 |
| cg09991769 | TRAF3 | 0,043 |
| cg12323063 | MAP2K6 | 0,043 |
| cg23044504 | NFKB2 | 0,044 |
| cg22819055 | TRAF3 | 0,044 |
| cg12970512 | TRAF3 | 0,046 |

**Table S6, related to the methods.** Homebrew Simoa assays antibodies, standards and running condition details.

**Table S5, related to Figure 6.** Probes differentially methylated between the low responders and the responders.
| Homebrew assay | Capture antibody | Detector antibody | Standard | Optimized conditions | Limit of detection |
| --- | --- | --- | --- | --- | --- |
| Pan-IFNα | ImmunoQure 8H1 | ImmunoQure 12H5 | PBL ref. 11150 | 2 steps<br>Detector 0,3 ug/ml<br>SBG 150pM | 0.05 fg/ml |
| IFNβ | PBL ref. 710322-9 | PBL ref. 710323-9 | PBL ref. 11415-1 | 2 steps<br>Detector 1 ug/ml<br>SBG 50pM | 0.07 pg/ml |
| IFNγ | Biolegend ref. 507501 | R&D ref. MAB285 | PBL ref. 11500 | 2 steps<br>Detector 0,3 ug/ml<br>SBG 150pM | 0.005 pg/ml |

